# Screening Problematic Internet Use in Parkinson’s disease: prevalence and clinical characterization

**DOI:** 10.64898/2026.07.21.26357985

**Authors:** Antônio Augusto Moreira Cardoso, Bruna Rafaela dos Santos Brito, André Vitor de Souza Fernandes, André Luiz de Souza Rodrigues, Bruno Lopes Santos-Lobato

## Abstract

**Background:** Problematic Internet use (PIU) has been scarcely investigated in Parkinson’s disease (PD). Objectives: To estimate the prevalence of PIU symptoms in PD using a specific screening instrument and to explore the clinical characteristics of these individuals.

**Methods:** Two-stage observational study in a Movement Disorders clinic. In Evaluation 1, participants with PD were screened by the Brazilian Portuguese Nine-Item Problematic Internet Use Questionnaire–Short Form (BR-PIUQ-SF-9). In Evaluation 2, screening-positive participants underwent an exploratory assessment of Internet use.

**Results:** 16.7% of participants with PD were positive for the BR-PIUQ-SF-9. The median Internet use time among these positive-screened participants was 5.5 hours per day. Most of them had impulsive-compulsive behaviors, and somatic concern, anxiety, and depressive mood were common psychiatric symptoms.

**Conclusions:** Approximately one in six participants screened positive for PIU symptoms. Impulsive-compulsive behaviors and depressive symptoms were frequent among those undergoing subsequent exploratory assessment.

---

Parkinson’s disease (PD) is the second most common neurodegenerative disease worldwide. Beyond classic motor symptoms, non-motor symptoms are common and impose a progressive burden on patients and their families [1]. Impulsive-compulsive behaviors (ICB), such as pathological gambling, hypersexuality, compulsive or binge eating, and compulsive buying, occur in 3.5 to 35% of people with PD, with a 5-year cumulative incidence of 46.1% and increase caregiver burden, representing challenges for PD management [2].

With the rise of the Internet and social media, these have become essential in daily life, regardless of socioeconomic conditions or age [3–5]. For people with PD, digital technologies may help compensate for reduced mobility and social participation, facilitating independence and social connection [6]. There is growing concern about Problematic Internet Use (PIU), which can be defined as excessive Internet use that causes a loss of sense of time while online and a growing need to be connected more often and for longer periods, at the expense of other aspects of their offline lives [7].

Evidence regarding PIU in PD remains limited. Previous studies have examined the PIU issue in the context of ICB, and the prevalence of PIU in PD remains poorly characterized. The present study aims to estimate the prevalence of PIU symptoms in PD using a specific screening instrument and to explore the clinical characteristics of these individuals.

## Methods

### Study design and ethical approval

We conducted an observational, two-stage study consisting of cross-sectional screening followed by an exploratory assessment of participants who screened positive. We recruited consecutive individuals with PD according to the UK PD Society Brain Bank clinical diagnostic criteria from the Movement Disorders Clinic at Hospital Ophir Loyola (Belém, Pará, Brazil). We enrolled participants between April 2023 and December 2025. The study was approved by the Hospital Ophir Loyola Ethics Committee (Protocol Number 6.433.041), and all participants provided written informed consent.

### Clinical assessment

We divided the clinical assessment into two different moments. In Evaluation 1, PIU symptoms among people with PD were screened using the Brazilian Portuguese version of the Nine-Item Problematic Internet Use Questionnaire (BR-PIUQ-SF-9) [8]. This instrument is a short-form screening measure for PIU symptoms in adolescents and adults [9] that assesses three dimensions of PIU: obsession, neglect, and control disorder, through nine questions, and the total score ranges from 9 to 45. A total score of 22 or higher has been proposed to identify individuals at risk of PIU [9].

For Evaluation 2, we implemented a comprehensive clinical protocol to assess Internet use among individuals with PD who had a positive BR-PIUQ-SF-9 score. The protocol consisted of: (I) Mobile Phone Problem Usage Scale — MPPUS-27 [10], (II) Self-perception of Text-message Dependency Scale — STDS [11], (III) Ten-Item Internet Gaming Disorder Test — IGDT-10 [12]. Also, we applied the Brazilian version of the Questionnaire for Impulsive-Compulsive Disorders in Parkinson’s Disease (QUIP-RS) [13,14], the 15-Item Geriatric Depression Scale (GDS-15) [15], the 8-Item Parkinson’s Disease Questionnaire (PDQ-8) [16], and the Brief Psychiatric Rating Scale (BPRS) [17].

### Statistical analysis

To identify factors associated with a positive BR-PIUQ-SF-9 screening in people with PD, we performed logistic regression. For univariate regressions, we selected the following independent variables: sex, age at evaluation, age at PD onset, PD duration, family history of PD, levodopa equivalent daily dose (LEDD), and use of dopaminergic agonists. Variables with associations with p < 0.1 were selected for multivariate logistic regression analyses. Comparisons of clinical data between screening-positive and screening-negative groups were performed using the chi-square and Mann-Whitney tests. Missing data were not imputed. All analyses were performed using SPSS for Windows version 23.0 (SPSS Inc., Chicago, USA).

## Results

We recruited 96 participants for this study who met the inclusion and exclusion criteria. Data are summarized in Table 1. The overall median and maximum scores in the BR-PIUQ-SF-9 questionnaire were 12 (IQR 9–20) and 25 points, respectively, and 33.3% of participants had the minimum score. Sixteen individuals with PD had a positive BR-PIUQ-SF-9 screening at Evaluation 1 (16.7%), with a median score of 22 (IQR 22-23). The most affected dimension was neglect. The frequency of people with a family history of PD was lower, and LEDD was higher in people with PD and PIU. Univariate logistic regression analyses showed that the tested independent variables were not associated with a positive BR-PIUQ-SF-9 screening in people with PD.

**Table 1.** Clinical characteristics of participants according to the BR-PIUQ-SF-9 screening.

| Variable | Available sample size | BR-PIUQ-SF-9 positive (n = 16) | BR-PIUQ-SF-9 negative (n = 80) | P-value |
| --- | --- | --- | --- | --- |
| Male sex, % (n) | 96 | 75 (12) | 62.5 (50) | 0.403 |
| Age at evaluation<sup>a</sup> | 94 | 60 (55-71) | 61.5 (55-69) | 0.698 |
| Age at onset<sup>a</sup> | 90 | 50 (40-65) | 50.5 (42-59) | 0.849 |
| Family history of PD, % (n) | 74 | 6.7 (1) | 20.3 (12) | **0.001** |
| Disease duration (years)<sup>a</sup> | 90 | 10 (9-13) | 8.5 (7-15) | 0.304 |
| Use of dopaminergic agonists, % (n) | 88 | 64.3 (9) | 38.6 (22) | 0.076 |
| Levodopa equivalent daily dose (mg/day)<sup>a</sup> | 89 | 975 (650-1233) | 750 (500-1012) | **0.045** |
| Hoehn & Yahr stage $\leq 2$, % (n) | 73 | 66.7 (10) | 84.5 (49) | 0.439 |
| BR-PIUQ-SF-9<sup>a</sup> | 96 | 22 (22-23) | 11 (9-19) | **<0.001** |
| BR-PIUQ-SF-9 Neglect dimension<sup>a</sup> | 96 | 2.67 (2.3-3) | 1.33 (1-2) | **<0.001** |
| BR-PIUQ-SF-9 Obsession dimension<sup>a</sup> | 96 | 2.5 (2.3-2.6) | 1 (1-2) | **<0.001** |
| BR-PIUQ-SF-9 Control dimension<sup>a</sup> | 96 | 2.33 (2-2.6) | 1 (1-2) | **<0.001** |
| Education (years)<sup>a</sup> | 74 | 11 (4-12) | 9 (5-11) | 0.254 |

<sup>a</sup> Values are presented as median (interquartile range). Available sample sizes vary due to missing data. Bold numbers indicate a p-value < 0.05
Abbreviations: BR-PIUQ-SF-9, Brazilian Portuguese version of the Nine-Item Problematic Internet Use Questionnaire; PD, Parkinson's disease.

Of the 16 individuals with PD who were positive on the BR-PIUQ-SF-9 at Evaluation 1, we evaluated 14 participants (two participants did not agree to be assessed in Evaluation 2). At Evaluation 2, the median MPPUS-27, STDS, and IGDT-10 scores were 101 (IQR 55-119), 27 (IQR 21-31), and 0, respectively. Thirteen individuals reported smartphone use, and the median daily time spent on smartphones and the Internet was 5.5 hours (IQR 3-8). Seven individuals (50%) reported using smartphones six hours or more per day. WhatsApp was the predominant messaging platform (85.7%). Median GDS-15 and PDQ-8 scores were 6.5 (IQR 3-11) and 39 (IQR 22-60), respectively, with 57.1% screened positive for depressive symptoms.

According to the QUIP-RS, five individuals exhibited ICB in a single domain, and five in multiple domains. Dopamine dysregulation syndrome was the only positive domain among participants with single-domain positivity. In persons with ICB across multiple domains, hobbyism/punding (80%) and dopamine dysregulation syndrome (60%) were the most affected domains (Fig. 1). The median BPRS score was 8.5 (IQR 7-16). The most frequently affected symptoms were somatic concerns, anxiety, and depressive mood. In contrast, psychotic symptoms were absent or uncommon.

**Fig. 1.**
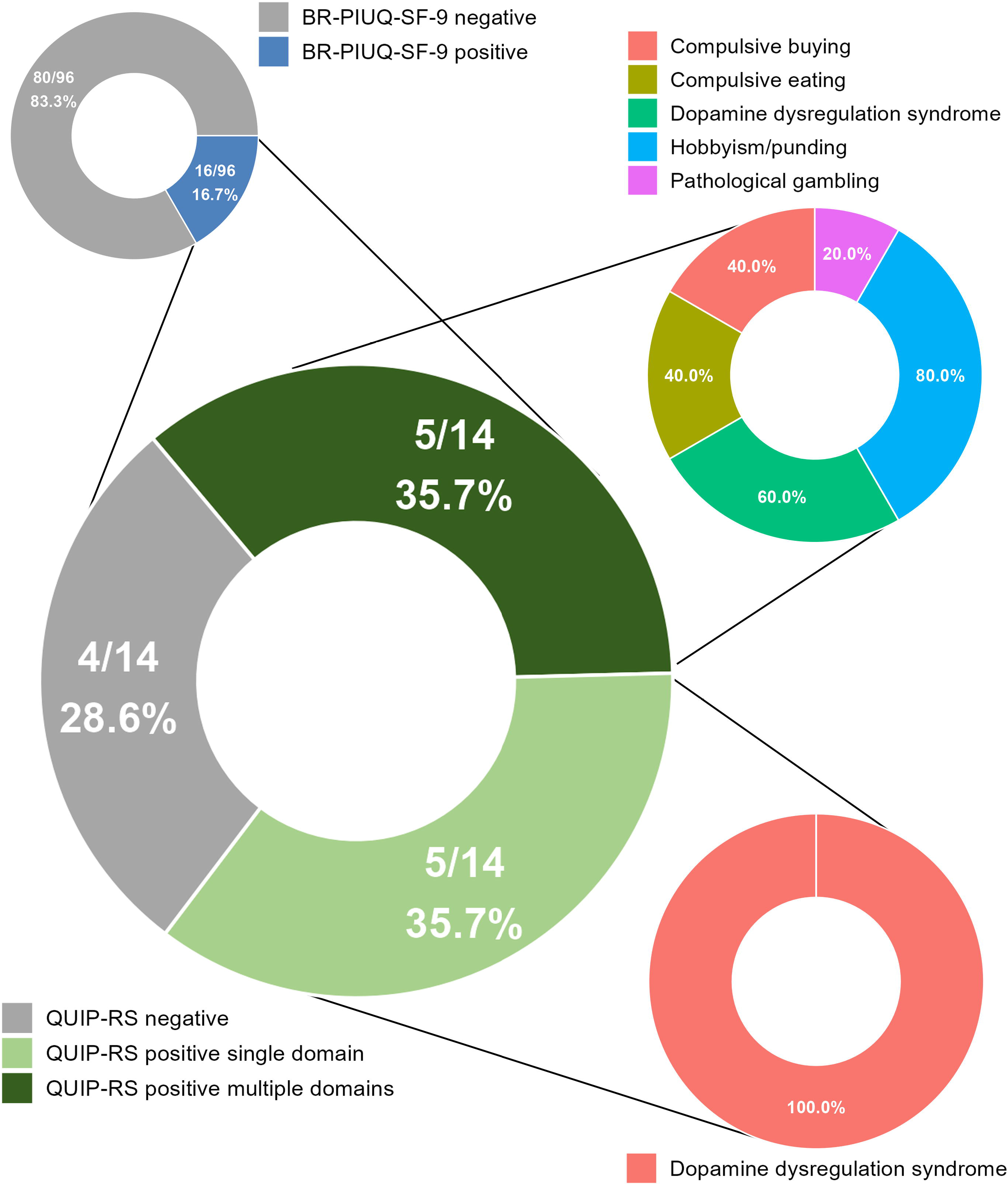
Distribution of impulsive-compulsive behaviors in participants with BR-PIUQ-SF-9 positive screening. The QUIP-RS was administered to a total of 14 participants among 96 individuals with PD, distributed across individuals with impulsive-compulsive behaviors in single or multiple domains. Dopamine dysregulation syndrome was present in all participants with single-domain positivity (light green). For QUIP-RS positive multiple domains (dark green), percentages are not mutually exclusive because participants could screen positive in more than one QUIP-RS domain. Abbreviations: BR-PIUQ-SF-9, Brazilian Portuguese version of the Nine-Item Problematic Internet Use Questionnaire; QUIP-RS, Brazilian Portuguese version of the Questionnaire for Impulsive-Compulsive Disorders in Parkinson’s Disease.

## Discussion

Using the BR-PIUQ-SF-9, we found that approximately one in six participants screened positive for PIU in our sample of people with PD. The psychiatric profile of people with PD and positive BR-PIUQ-SF-9 was characterized mainly by somatic concern, anxiety, and depressive mood, with substantial variability in quality of life. ICB were observed in these individuals, with dopamine dysregulation syndrome being the most frequent positive domain. The Internet was accessed primarily via messaging platforms on smartphones for approximately one-quarter of the day, whereas electronic gaming was uncommon.

The presence of PIU in PD has generally been examined in the context of ICB. In China, “compulsive Internet browsing” was detected in only one participant from 312 individuals with PD [18], and case reports showed that people with PD could use the Internet for pathological gambling [19,20]. A previous study found higher scores on an Internet-adapted scale of obsessive-compulsive symptoms among selected patients with PD and ICB than among patients without ICB and healthy controls [6]. So far, despite PIU being recognized as an abnormal behavior in the context of PD, little is known about its prevalence.

In another study, which applied an 18-item version of the Problematic Internet Use Questionnaire to regular Internet users with PD, it was reported that incorporating the questionnaire increased the recognition of ICB, particularly pathological gambling and compulsive sexual behavior [21]. Their findings support the possibility that PIU symptoms may serve as a clinical marker of ICB. In our study, 10 of the 14 participants previously screened positive for BR-PIUQ-SF-9 also screened positive in at least one QUIP-RS domain at Evaluation 2. However, our findings on the association between PIU and ICB should be interpreted with caution compared with previous studies, as our QUIP-RS assessment was administered only to screening-positive participants and at a later point.

Detecting PIU has been challenging, and while multiple tools assess the condition, none is considered a gold standard [8]. Also, most technological addiction disorders have been explored in children, adolescents, and young adults [22]. Thus, screening instruments such as the BR-PIUQ-SF-9 are not diagnostic tools for PIU [9].

When validated in Brazil, the BR-PIUQ-SF-9 demonstrated reliable psychometric properties, with good internal consistency and moderate test-retest reliability [8]. Compared with the present study, the validation study recruited participants with a mean age of 38 years. Despite reporting that only 4.2% of participants in the validation study had the minimum score for the instrument, the larger floor effect in our sample suggests that many participants endorsed no PIU symptoms, which may reflect lack of insight, lower exposure to digital technologies, differences in item interpretation, or limited sensitivity of the instrument in this population [23].

Elderly populations worldwide are spending more time on the Internet. A Korean study on smartphone use in 3,121 individuals over 60 years showed that most participants reported two hours or less of use, and 14.6% of participants were classified in the smartphone addiction risk group, and that smartphone usage time was positively related to smartphone addiction [24]. In an analysis of more than 200,000 adults in Brazil, individuals aged 60 years or older reported increased use of smartphones, computers, and tablets for leisure over a period of six years [5]. In the present study, half of the positive BR-PIUQ-SF-9 individuals reported spending 6 hours or more of daily Internet access, while a previous report estimated an average of 9 hours and 9 minutes per day among Brazilian Internet users aged 16 years or older [25].

As limitations, the present study recruited a small sample at a single center with a low number of positive-screened events, which limits the generalizability of the results. The BR-PIUQ-SF-9 and its cut-off values, as well as all other specific Internet use instruments, were not validated in elderly individuals nor in people with PD. Also, only screening-positive participants were included in the clinical protocol for assessing Internet use, and the interval between the two evaluations was not standardized and could not be determined. Therefore, clinical, psychiatric, and behavioral measures obtained during the second stage may not reflect the participants’ status at the time of the initial PIU screening. The absence of an age- and sex-matched healthy control group limits the interpretation of the results.

In conclusion, the BR-PIUQ-SF-9 was feasible for screening PIU symptoms in people with PD. However, its diagnostic accuracy in PD remains to be established. Our sample presented a prevalence of 16.7% of positive BR-PIUQ-SF individuals with PD, many of them with ICB.

Considering the increased access to the Internet among elderly people and individuals with PD, the possible overlap between PIU and ICB, and the social impacts of this issue, further studies are needed to explore this theme.

## Data Availability

All data produced in the present study are available upon reasonable request to the authors.

## Author Roles

(1) Research project: A. Conception, B. Organization, C. Execution; (2) Statistical Analysis: A. Design, B. Execution, C. Review and Critique; (3) Manuscript: A. Writing of the first draft, B. Review and Critique

A.A.M.C: 1A, 1C, 2A, 2C, 3B.

B.R.S.B.: 1A, 1C, 2A, 2C, 3B.

A.V.S.F.: 1C, 2C, 3B.

A.L.S.R.: 1C, 2C, 3B.

B.L.S-L.: 1A, 1B, 1C, 2A, 2B, 2C, 3A, 3B.

## Acknowledgements

We thank Dr. Pedro Melo Barbosa for reviewing the manuscript and suggesting insightful contributions.

## Disclosures

### Ethical Compliance Statement

The study was approved by the Hospital Ophir Loyola Ethics Committee (Protocol Number 6.433.041). All participants provided written informed consent. We confirm that we have read the Journal’s position on issues involved in ethical publication and affirm that this work is consistent with those guidelines.

### Financial Disclosures for the previous 12 months

B.L.S-L. reports a research grant funded by the Brazilian National Council for Scientific and Technological Development.

### Data Availability Statement

The datasets generated during and/or analyzed during the current study are available from the corresponding author on reasonable request.

## References

[1] T. Foltynie, V. Bruno, S. Fox, A.A. Kühn, F. Lindop, A.J. Lees, Medical, surgical, and physical treatments for Parkinson’s disease, Lancet 403 (2024) 305–324.

[2] A.H. Evans, D. Okai, D. Weintraub, S.-Y. Lim, S.S. O’Sullivan, V. Voon, P. Krack, C. Sampaio, B. Post, A.F.G. Leentjens, P. Martinez-Martin, G.T. Stebbins, C.G. Goetz, A. Schrag, Members of the International Parkinson and Movement Disorder Society (IPMDS) Rating Scales Review Committee, Scales to assess impulsive and compulsive behaviors in Parkinson’s disease: Critique and recommendations, Mov Disord 34 (2019) 791–798.

[3] L. Yang, C. Cao, E.D. Kantor, L.H. Nguyen, X. Zheng, Y. Park, E.L. Giovannucci, C.E. Matthews, G.A. Colditz, Y. Cao, Trends in Sedentary Behavior Among the US Population, 2001-2016, JAMA 321 (2019) 1587–1597.

[4] S.A. Prince, A. Melvin, K.C. Roberts, G.P. Butler, W. Thompson, Sedentary behaviour surveillance in Canada: trends, challenges and lessons learned, Int J Behav Nutr Phys Act 17 (2020) 34.

[5] P.C. Cardoso, T.C.M. Caldeira, T.M. de Sousa, R.M. Claro, Changes in Screen Time in Brazil: A Time-Series Analysis 2016-2021, Am J Health Promot 37 (2023) 681–684.

[6] K. Wu, M. Politis, S.S. O’Sullivan, A.D. Lawrence, S. Warsi, A. Lees, P. Piccini, Problematic Internet use in Parkinson’s disease, Parkinsonism Relat Disord 20 (2014) 482–487.

[7] C.L. Lortie, M.J. Guitton, Internet addiction assessment tools: dimensional structure and methodological status, Addiction 108 (2013) 1207–1216.

[8] D.T. Spritzer, W. de L. Machado, M.B. Yates, V.R. Astolfi, P. Laskoski, C. Pessi, S. Laconi, K. Kaliszewska-Czeremska, Z. Demetrovics, O. Király, I.C. Passos, S. Hauck, Psychometric Properties of the Nine-Item Problematic Internet Use Questionnaire in a Brazilian General Population Sample, Front Psychiatry 12 (2021) 660186.

[9] B. Koronczai, R. Urbán, G. Kökönyei, B. Paksi, K. Papp, B. Kun, P. Arnold, J. Kállai, Z. Demetrovics, Confirmation of the three-factor model of problematic internet use on off-line adolescent and adult samples, Cyberpsychol Behav Soc Netw 14 (2011) 657–664.

[10] A. Bianchi, J.G. Phillips, Psychological predictors of problem mobile phone use, Cyberpsychol Behav 8 (2005) 39–51.

[11] T. Igarashi, T. Motoyoshi, J. Takai, T. Yoshida, No mobile, no life: Self-perception and text-message dependency among Japanese high school students, Comput. Human Behav. 24 (2008) 2311–2324.

[12] O. Király, P. Sleczka, H.M. Pontes, R. Urbán, M.D. Griffiths, Z. Demetrovics, Validation of the Ten-Item Internet Gaming Disorder Test (IGDT-10) and evaluation of the nine DSM-5 Internet Gaming Disorder criteria, Addict Behav 64 (2017) 253–260.

[13] D.F. Guerra, A.E. Lemos Silva, T. da S.R. Paz, L.N.S. Filho, L.F. Vasconcellos, V.L.S. de Britto, S. Allodi, D. Weintraub, A. Swarowsky, C.L. Correa, Measurement properties from the Brazilian Portuguese version of the QUIP-RS, NPJ Parkinsons Dis 6 (2020) 6.

[14] D. Weintraub, E. Mamikonyan, K. Papay, J.A. Shea, S.X. Xie, A. Siderowf, Questionnaire for Impulsive-Compulsive Disorders in Parkinson’s Disease-Rating Scale, Mov Disord 27 (2012) 242–247.

[15] D. Weintraub, K. Saboe, M.B. Stern, Effect of age on geriatric depression scale performance in Parkinson’s disease, Mov Disord 22 (2007) 1331–1335.

[16] J. Dams, J. Klotsche, B. Bornschein, J.P. Reese, M. Balzer-Geldsetzer, Y. Winter, A. Schrag, A. Siderowf, W.H. Oertel, G. Deuschl, U. Siebert, R. Dodel, Mapping the EQ-5D index by UPDRS and PDQ-8 in patients with Parkinson’s disease, Health Qual Life Outcomes 11 (2013) 35.

[17] J.A.S. Crippa, R.F. Sanches, J.E.C. Hallak, S.R. Loureiro, A.W. Zuardi, Factor structure of Bech’s version of the Brief Psychiatric Rating Scale in Brazilian patients, Braz J Med Biol Res 35 (2002) 1209–1213.

[18] W. Fan, H. Ding, J. Ma, P. Chan, Impulse control disorders in Parkinson’s disease in a Chinese population, Neurosci Lett 465 (2009) 6–9.

[19] A.J. Larner, Medical hazards of the internet: gambling in Parkinson’s disease, Mov Disord 21 (2006) 1789.

[20] S.H. Wong, Z. Cowen, E.A. Allen, P.K. Newman, Internet gambling and other pathological gambling in Parkinson’s disease: a case series, Mov Disord 22 (2007) 591–593.

[21] M. Kovács, A. Makkos, D. Pintér, A. Juhász, G. Darnai, K. Karádi, J. Janszky, N. Kovács, Screening for Problematic Internet Use May Help Identify Impulse Control Disorders in Parkinson’s Disease, Behavioural Neurology 2019 (2019) 4925015.

[22] J. Sherer, P. Levounis, Technological Addictions, Psychiatr Clin North Am 45 (2022) 577–591.

[23] A. Avila, X. Cardona, J. Bello, P. Maho, F. Sastre, M. Martin-Baranera. Impulse control disorders and punding in Parkinson’s disease: the need for a structured interview, Neurologia 26 (2011) 166–172.

[24] J.-H. Jeong, S.-M. Bae, The Relationship Between Types of Smartphone Use, Digital Literacy, and Smartphone Addiction in the Elderly, Psychiatry Investig 19 (2022) 832–839.

[25] We Are Social, Meltwater. Digital 2025: Global Overview Report. 2025. Available at: https://wearesocial.com/wp-content/uploads/2025/02/GDR-2025-v2.pdf. Accessed July 5, 2026.

